# COVID-19 pandemic-related lockdown: response time is more important than its strictness

**DOI:** 10.1101/2020.06.11.20128520

**Authors:** Gil Loewenthal, Shiran Abadi, Oren Avram, Keren Halabi, Noa Ecker, Natan Nagar, Itay Mayrose, Tal Pupko

## Abstract

The rapid spread of SARS-CoV-2 and its threat to health systems worldwide have led governments to take acute actions to enforce social distancing. Previous studies used complex epidemiological models to quantify the effect of lockdown policies on infection rates. However, these rely on prior assumptions or on official regulations. Here, we use country-specific reports of daily mobility from people cellular usage to model social distancing. Our data-driven model enabled the extraction of mobility characteristics which were crossed with observed mortality rates to show that: (1) the time at which social distancing was initiated is of utmost importance and explains 62% of the number of deaths, while the lockdown strictness or its duration are not as informative; (2) a delay of 7.49 days in initiating social distancing would double the number of deaths; and (3) the expected time from infection to fatality is 25.75 days and significantly varies among countries.

## Introduction

In 2020, the coronavirus pandemic has rapidly spread around the globe, threatening health and economical systems. At first, many governments attempted to minimize exposure to the virus by limiting cross-border arrivals. However, the rapid person-to-person transmission rate of the virus^1,2^ required that more severe measures be taken to plummet infection frequencies. Governments that used lockdown to enforce social distancing varied in their policy, timing, and duration, in particular relative to the mortality rate in their country. For example, Italy enforced a severe, nationwide, lockdown on March 10, when over 35,000 confirmed cases and almost 3,000 deaths had already been recorded. In other countries, lockdown policies were embraced at earlier stages in attempt to prevent severe outbreaks. Israel, for instance, reached the strict lockdown on March 19 with a relatively low number of 648 confirmed cases and no deaths to that day. In contrast, several countries, such as Sweden and Japan, advocated social distancing but did not enforce a lockdown as a means of coronavirus spread prevention.

How can social distancing be quantified? One could measure governmental regulations such as the permitted walking distance from the residence, limitations on mass gatherings, school closures, and whether people were allowed to attend their workplaces. For example, Hu et al. suggested a score that takes into account various governmental interventions in the United States^3^. This score was used to predict future infections depending on the intervention level. While this model may be useful when governmental decisions are made, it does not reflect whether social distancing has been implemented *de facto*^4^. Soures et al. used data collected from navigation applications on mobile cellphones together with past infection rates to predict future infection rates^5^. These predictions were based on a neural-network model, in which the connection between mobility data and infection rates is hard to interpret, and thus, practically, cannot be converted into tangible measures for the arms race against the disease. It is currently unknown which aspects of the lockdown (e.g., duration, strictness, timing from onset of death cases) affect mortality rates. Understanding the linkage between lockdown dynamics and COVID-19 death incidents is highly important for balancing between health, welfare, and economy.

Here we develop parametric models that quantify trends related to mobility and mortality and fit them to all OECD countries. Using these models, we demonstrate that the timing in which the social distancing was initiated by itself explains 62% of the COVID-19 related deaths across OECD countries, excluding Japan (that was previously reported as an exception with respect to the spread of the disease, e.g., by Iwasaki and Grubaugh^6^). In contrast, the severity of the lockdown and its duration are not as informative for explaining mortality rates. Our analysis thus suggests that a moderate lockdown, rather than a very strict one as imposed by most countries, should be sufficient to curb COVID-19 related mortality, as long as action is taken in the appropriate time frame.

## Results

Following the COVID-19 outbreak, Apple Inc. published daily reports regarding people movement, collected from usage of maps on mobile cellphones^7^. We used these mobility data, denoted as *M*(*t*), to quantify the actual commencement of the lockdown as a function of time in different OECD countries. We collected daily death incidents across time and overlaid them on the mobility data. We observed that the trend of daily deaths stabilized and subsequently decreased several days after a sharp mobility drop (Fig. 1a for the United Kingdom as an example and Supplementary Fig. S1 for all OECD countries).

**Figure 1.**
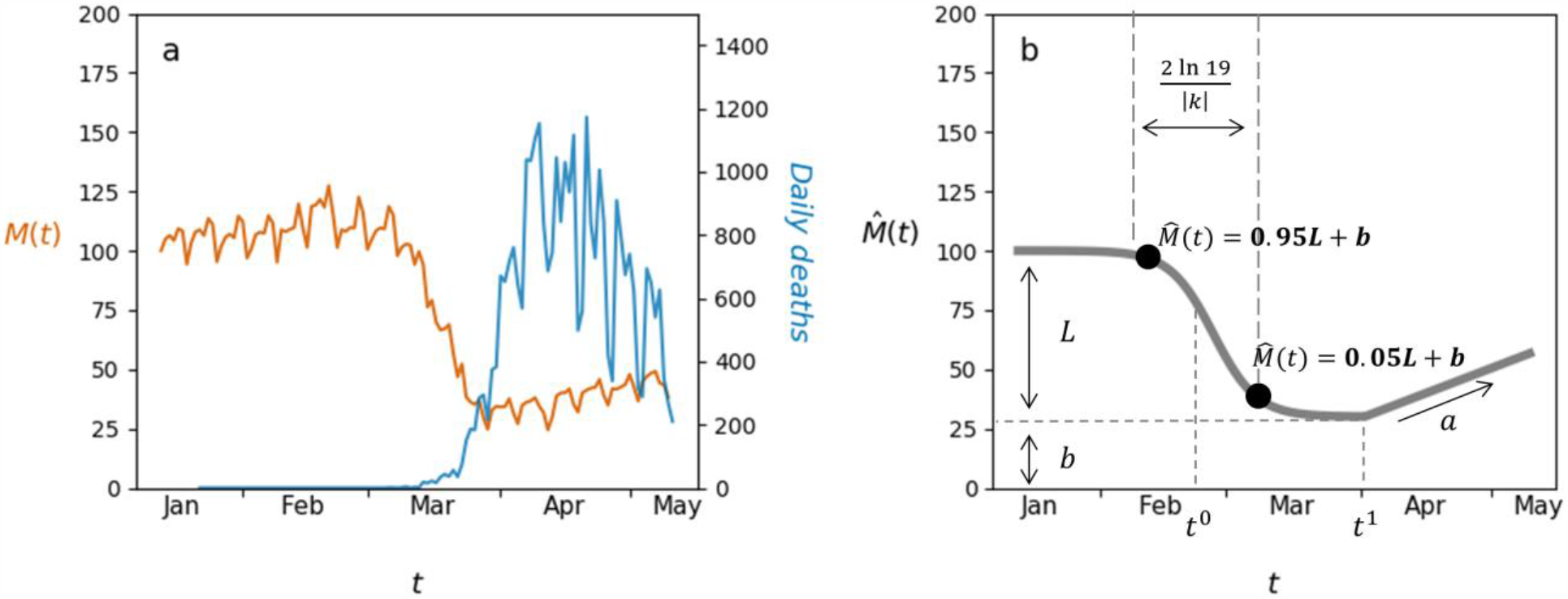
Modeling mobility data. (a) Daily mobility data, *M*(*t*), (orange line, left y axis) overlaid with daily deaths (blue line, right y axis) for the United Kingdom. *M*(*t*) is given as percentages relative to that recorded on January 13, that serves as the baseline; (b) An illustration of the mobility model and its free parameters: *L*-mobility difference between routine and lockdown; - drop steepness; *t*^0^-drop midpoint; - mobility level during lockdown; *t*^1^-release day; *a*-recovery rate, see Methods for more details.

### Mobility analysis

The mobility in a single country during the observed time period of the COVID-19 pandemic can generally be divided into four phases: (1) a stable phase of high mobility (with fluctuations on weekends); (2) a sharp drop (suggesting social distancing has actually started); (3) a period of low mobility; and (4) a gradual incline towards a normal routine (Fig. 1). Phases (1)-(3) reflect a (mirrored) logistic function and phase (4) is approximately linear. We modeled this overall trend by assembling a logistic function and a linear one as a function of time (*t*, given in days):

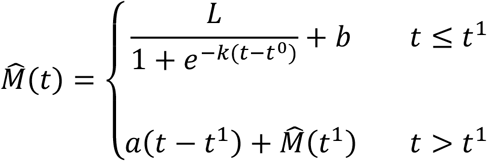

The six free parameters of this model are illustrated in Fig. 1b.

Fitting the mobility model to the 37 OECD countries resulted in an average Pearson correlation of *r*^*2*^ = 0.9 between the observed data and the fitted functions (all *P values* < 10^−32^, Supplementary Table S1). The inferred model parameters enabled the comparison of several informative features for the different countries (see Methods). As examples, we present the fitted models for five representative OECD countries: Germany, Israel, Italy, Spain, and Sweden (Fig. 2; for the inferred features and fitted models of all countries see Supplementary Table S2 and Supplementary Fig. S2). Our results demonstrate that while the *lockdown strictness* varied considerably, all countries reached some form of a lockdown by the middle of March 2020, with Spain presenting the most intense drop (88%). The *social distancing start time* in Italy occurred earlier, on February 25 compared to March 6-9 for the abovementioned four other countries. Nevertheless, the mobility in Italy declined in a relatively gradual manner with respect to other examined countries, as the *drop duration* lasted 20 days. The extent of mobility reduction in Germany (59%) was relatively low compared to other countries in which a lockdown was issued, and a gradual return to normal routine was initiated right after the lowest mobility level was reached. Even though a lockdown was not regulated in Sweden, the data and model show that social distancing indeed happened, as a drop of 29% was observed followed by a moderate return back to routine (*lockdown release rate* of 0.57).

**Figure 2.**
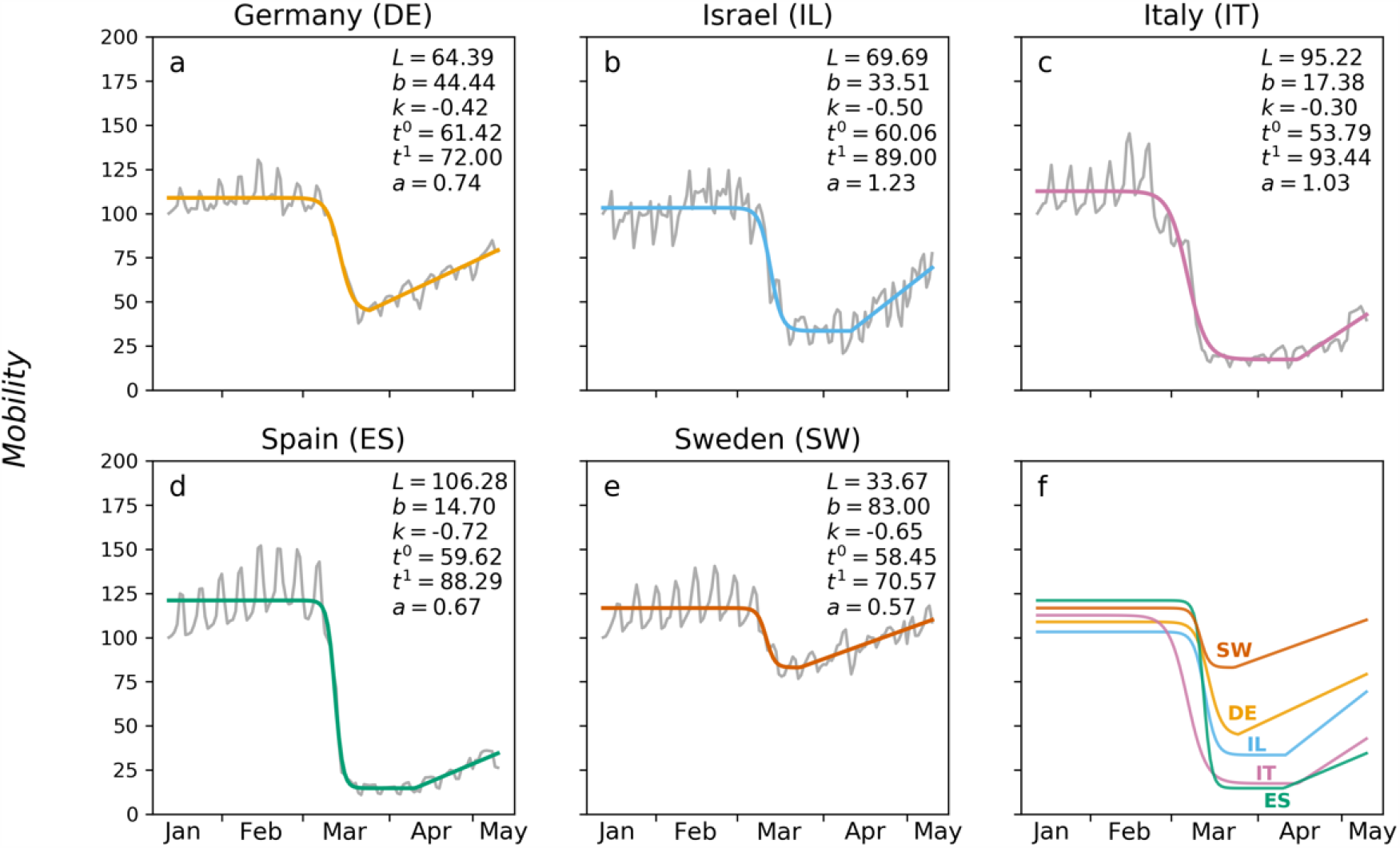
The fit of the mobility model for five representative OECD countries. Colored lines in panels a-e represent the mobility model 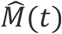 fitted to the mobility data *M*(*t*) (grey lines). The optimized parameters are indicated. Panel f presents the overlay of the five fitted models. The two-letter codes and the five colors correspond to the countries represented in panels a-e (countries abbreviations are denoted in the titles of the panels). The x axes represent days from January 13 to May 10, 2020. The y axes represent the percentage change in mobility. For the parameter values and the inferred features of all 37 countries, see Supplementary Tables S1-S2.

### COVID-19 mortality

The accumulated number of COVID-19 deaths across time, *D*(*t*), was fitted using a logistic function, 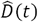, to the data (see Methods). The average Pearson correlation between the observed mortality data and the fitted function, across all countries, was *r*^2^ = 0.99 (all *P values* < 1*e* − 98; Supplementary Table S3; see Fig. 3 for examples of Israel and Japan and Supplementary Fig. S2 for all countries). From the fitted model we computed the *COVID-19 Mortality Probability* for each country (see Methods).

**Figure 3.**
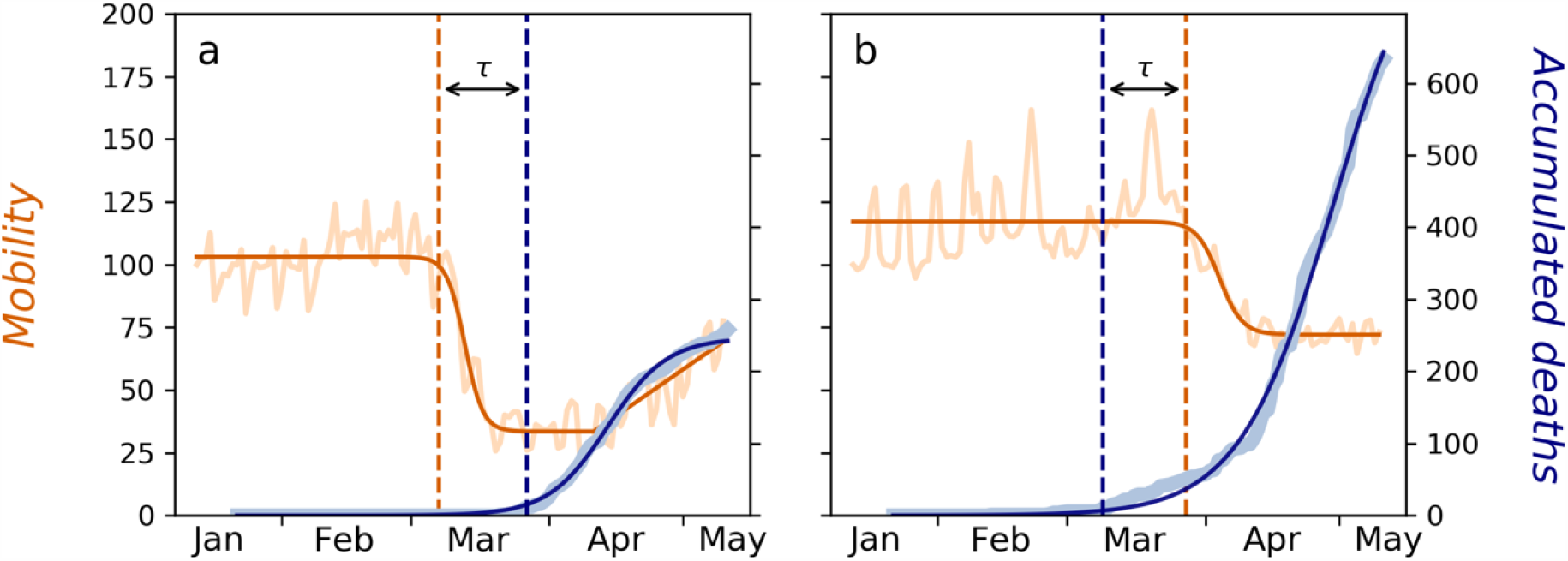
Synchronizing between the death incidents model and the mobility model. The dark orange plots represent the mobility model, 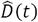, fitted to the mobility data, *M*(*t*) (light orange; left y axis) of (a) Israel and (b) Japan. The dashed vertical orange line represents the *social distancing start time*, predicted by the mobility model. The dark blue plots represent the death model, 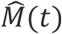, fitted to the accumulated death data, *D*(*t*) (light blue; right y axis). The dashed vertical blue lines represent the day ten deaths were documented. *τ* represents the time difference between the orange and the blue vertical lines, defined as the response time (*τ* is negative for Israel and positive for Japan). The graphs for all OECD countries are given in Supplementary Fig. S2.

### Association between mobility and mortality data

We computed the response time of each country, *τ*, defined as the difference between the *social distancing start time* and the day when ten first deaths were recorded. Figure 3 demonstrates the computation of *τ* for Israel and Japan (*τ* = − 19.83 and 18.68 days, respectively; see Supplementary Fig. S2 and Supplementary Table S4 for all countries). While a negative *τ* was inferred for most countries, a positive *τ* was inferred for five countries (France, Italy, Japan, Spain, and the United States), indicating that social distancing started after ten COVID-19 deaths were documented (Fig. 4, Supplementary Fig. S2). We observed a significant correlation between *τ* and the log *COVID-19 Mortality Probability* (Pearson *r*^2^ = 0.36, *P values* < 1*e* − 4). Previous reports have discussed the abnormally low mortality rate in Japan^6^, thus, we computed the correlation excluding Japan and obtained a substantial increase in correlation (*r*^2^ = 0.6^2^, *P values* < 1*e* − 8). Neither the *lockdown strictness* nor the *lockdown duration* were significantly correlated with log *COVID-19 Mortality Probability* (Supplementary Table S5).

**Figure 4.**
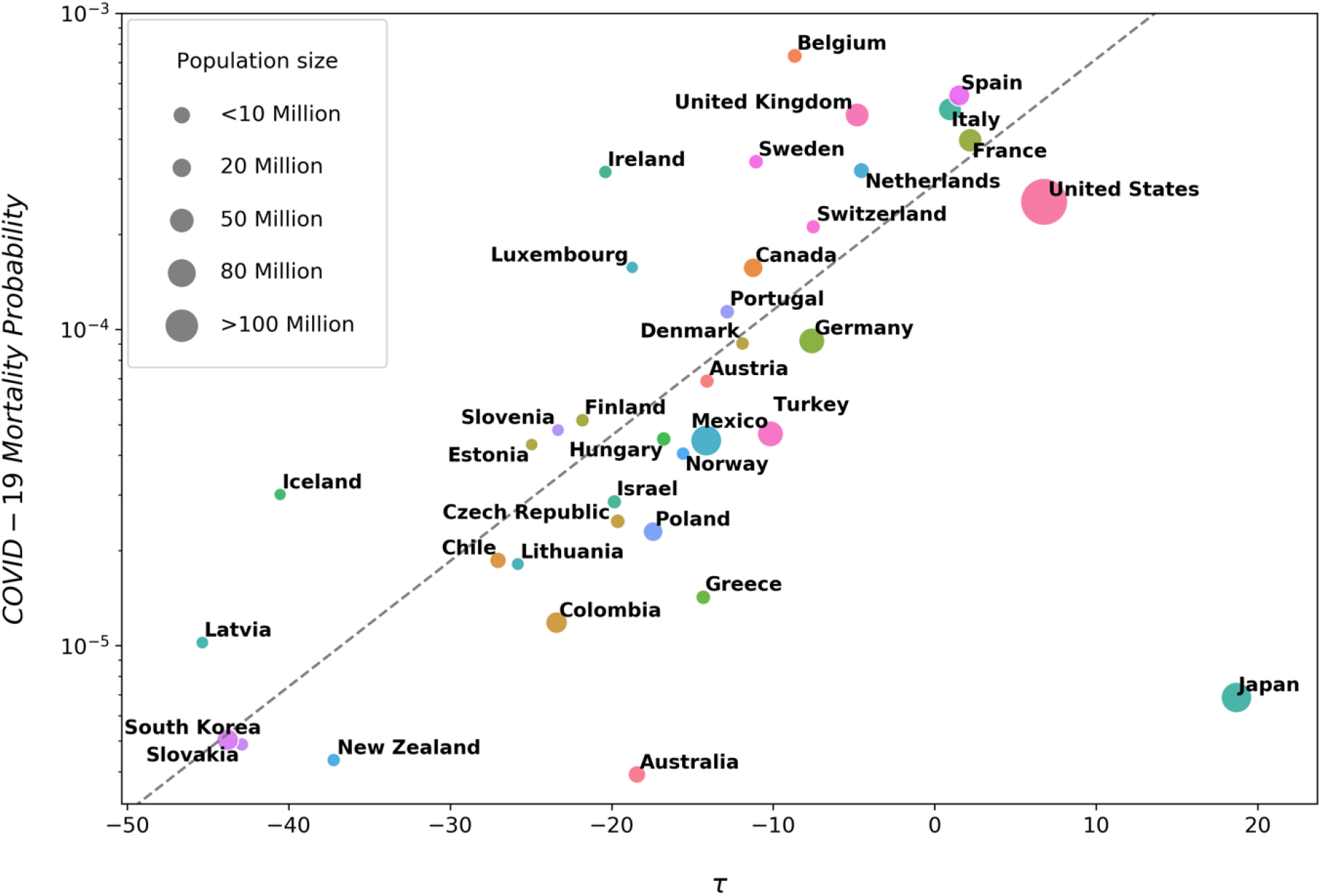
A semi-logarithmic scatter plot of the *COVID-19 Mortality Probability* and *τ*. The x axis represents *τ*, the difference between the *social distancing start time* and the day in which the ten first deaths were recorded for the respective country (intuitively, the response time). The y axis represents the *COVID-19 Mortality Probability* in a logarithmic scale. Dot sizes are proportional to population sizes. The dashed line corresponds to the fitted regression, excluding Japan. For raw data, see Supplementary Table S4.

The high correlation between *τ* and the log *COVID-19 Mortality Probability* yielded a crucial implication, as it allowed inferring the time required for this probability to double. We fitted a linear regression to the data presented in Fig. 4 (excluding Japan) and computed the mortality doubling time according to the slope of the fitted regression line. Accordingly, our results indicate that a 7.49 days delay in lockdown commencement doubles the expected number of deaths (95% CI 6.0^2^, 10.03]; see Methods). This result is in accordance with two previous studies. First, using empirical epidemiological study in Wuhan city, China, Li et al. estimated that the infection doubling time was 7.4 days^2^. Second, using epidemiological models, Pei et al. estimated that 55% of the deaths could have been avoided if non-pharmaceutical interventions had been implemented a week earlier^8^.

Next, we aimed to compute the time offset between mobility reduction and the decline in mortality growth rate. This offset serves as a proxy for the time between infection and fatality. The time offset that yielded the highest correlation between *M*(*t*) and mortality growth rate was 25.75 days on average across countries (average Pearson *r*^2^ = 0.49, all *P values* < 0.004 except for Iceland, Latvia, and Slovakia, which had only several death incidents; Supplementary Table S6 and Supplementary Fig. S3). This result highly resembles previous estimations of the time from infection to fatality^9,10^, although here it was derived from entirely different data and analysis. Interestingly, this offset highly varied among the countries (*σ* = 7.98 days), with exceptionally low offset in the United Kingdom (eight days) and exceptionally high offset in Finland, Hungary, and Estonia (38-39 days). There was a mild correlation between the offset and *COVID-19 Mortality Probability* (*r*^2^ = 0.34), namely, the time between exposure and fatality shortens as the mortality rate grows.

## Discussion

In this study, we modeled the mobility dynamics across time during the COVID-19 pandemic. Using this model we computed explanatory features that characterize a lockdown, and in turn, these features provided a quantitative measure for comparing the lockdown dynamics and outcome across countries. We found high correlation between the response time of a country and its mortality rate. This finding suggests that countries that took early measures to limit population mixing had better control on the viral-related mortality. In contrast, neither the *lockdown duration* nor the *lockdown strictness* were significantly associated with mortality rates (Supplementary Table S5). These results imply that a tight lockdown could have been unnecessary.

Official governmental regulations are possible resources for extracting features regarding the lockdown. Such features were examined for their association with infection rates in previous studies^5,11^. However, the time governmental regulations were declared was often significantly different from the date social distancing initiated *de facto*^4^. For example, Italy announced regional lockdowns in three phases. Initially, a lockdown was applied in Lombardy and Veneto regions on February 20. This lockdown was expanded to all of Northern Italy on March 8, and finally to a nationwide lockdown on March 11. However, the mobility data show that even though the initial lockdown was declared on Lombardy, mobility volumes across Italy kept elevating and started to decrease only a week later. In contrast, when the lockdowns on Northern Italy and later, on the entire country, were declared, a large drop in mobility usage had already taken place (Supplementary Fig. S4).

Most of the examined OECD countries complied well to the regression analysis in this study. Small deviations could be explained by modest variations between countries, such as the conditions for defining a patient as a SARS-CoV-2 carrier, or by differences in mobile usage across areas. Notably, different geographical areas also differ in numerous other attributes that may affect the coronavirus spread and induced mortality rates, e.g., humidity, wind speed, ethnicity, viral genotypic variation, and cultural habits. One eminent example emerging from our analysis is Japan, where a relatively low mortality rate occurred even though mobility reduction took place relatively late. The low mortality rate in Japan has previously been discussed^6^, and it should be beneficial to better understand the different trajectory of the pandemic in Japan, with respect to the Japanese governmental regulations and customs as a possible alternative to a strict lockdown. Combining these features with the proposed mobility model may increase its overall accuracy.

The inferred high association between mobility and mortality rate led to crucial implications. First, we found that the number of death incidents would double if action was taken in a delay of approximately one week. Second, we inferred that the time offset from exposure to fatality is roughly 26 days, with some countries exhibiting extremely shorter time offsets and others much longer. We also observed that the correlation between this time offset and *COVID-19 Mortality Probability* results in *r* = −0.58. We hypothesize that the inverse correlation between the two may be explained by the load of high mortality on the health system, which might hamper the treatment and shorten survival time. The remaining variation may be explained by different treatment protocols, COVID-19 test policies, or other characteristics, and requires additional study.

We focused our analysis on 37 OECD countries, to concentrate on a representative group of relatively reliable reports. Nevertheless, our results sustain when including additional non-OECD countries or when concentrating on sub regions for which sufficient data exist: the Pearson *r*^2^ between *τ* and the log *COVID-19 Mortality Probability* for 58 countries was 0.36 (*P values* < 4*e* − 5; Supplementary Fig. S5). A significant correlation (*r*^2^ = 0.35) was also observed when analyzing states within the United States (*P values* < 1*e* − 5; Supplementary Fig. S6). Finally, while in this study we investigated the association of lockdown and mortality, the number of confirmed cases or hospital-committed patients can also be examined. We opted to use mortality as the outcome since it is less dependent on the number of COVID-19 tests, which widely varies among different countries. Nevertheless, significant Pearson *r*^2^ of 0.47 between *τ* and the log *COVID-19 Infection Probability* in OECD countries was also observed (*P values* < 1*e* − 5; Supplementary Fig. S7).

The results of our analysis show that social distancing is a major factor in controlling COVID-19 spread. Similar to previous studies^2,8^, our analysis shows that if social distancing is not adopted, death incidents are doubled every 7.49 days. However, it also shows that a strict lockdown policy is not required. Therefore, to avoid major infection outbreaks, we suggest undertaking a moderate form of a lockdown that can be tolerated by the society for longer time periods, with minimal socio-economic damage.

## Methods

### Mobility data

Mobility data, *M*(*t*), with one data point per day, *t*, were downloaded from Apple repository on May 11, 2020^7^. The Apple dataset reports the daily volume of population driving, walking, or using transit (public transportation) in a specified region, and is reported as the percentage with respect to a benchmark (100%) set on January 13th, 2020. Data for a total of 119 time points were downloaded, with *t*_*1*_ and *t*_*119*_ corresponding to January 13 and May 10, respectively. Due to the high similarity between “walking” and “driving” data (average Pearson correlation across countries *r*^2^ = 0.91, *σ* = 0.07, all *P values* < 10^−129^) and since the “transit” data are incomplete, all analyses were applied using the “driving” data only.

To fit 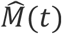 to the mobility data, *M*(*t*), and receive the values of the parameters for every country, we used the Levenberg–Marquardt optimization algorithm from the SciPy module^12–14^. According to the parameters inferred from 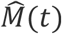, we computed seven features to characterize the mobility trend in a country. (1) *t*′, *Social distancing start time*; and (2) *t*″, *Minimal mobility time point*, corresponding to the times before and after the mobility drop. These points are defined as time 95% and 5% of the drop, parameterized by *L*, and *t* is the middle time point between them. Thus, 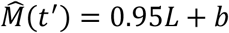 and 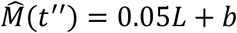. Then,

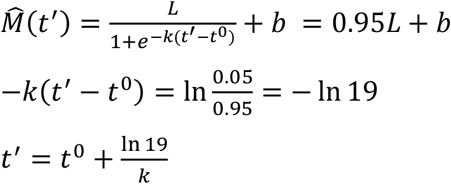

Similarly, 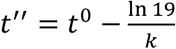 (note that *k* is negative); (3) *Drop duration*, the time difference 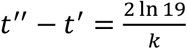; (4) *Lockdown release day t*; (5) *Lockdown strictness* 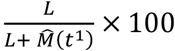, such that 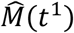 is the function value at the release day; (6) *Lockdown duration t*^−^*t*″; (7) *Lockdown release rate a* (the slope of the linear function).

### Mortality data

The daily cumulative numbers of COVID-19 related mortalities, *D*(*t*), were downloaded from the COVID-19 Data Repository by the Center for Systems Science and Engineering (CSSE) at Johns Hopkins University^15^. The data were available for the period of January 22 to May 10, 2020 (a total of 110 data points). The cumulative number of deaths in Australia, Canada, and the United States were aggregated across the regions reported in the dataset within each of these countries. A logistic function 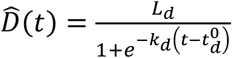 was fitted to *D*(*t*), as in Tátrai and Várallyay^16^, using the Levenberg–Marquardt optimization algorithm from the SciPy module^12–14^. The parameters *L*_*d*_,*k*_*d*_, and 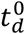 are similar to those defined for the mobility model and represent the total expected mortality at the end of the pandemic, mortality increase rate, and the time the cumulative mortality has reached its midpoint, respectively. To infer the *COVID-19 Mortality Probability*, the *expected mortality* (*L*_*d*_) was normalized by the population size for each country. Data of population size were obtained from the World Population Review website^17^. Of note, our goal in this work was not to predict mortality rates, but rather, to find correlates with large changes in mortality patterns across countries. Since we correlate with the logarithm of the mortality rates, we expect that small deviations in mortality estimates will not affect our conclusions.

### Association between mortality and response time

We define a response time, *τ*, as the difference between two time points: the *social distancing start time* (as inferred from 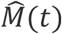) and the day in which ten first COVID-19 related deaths were recorded (according to *D*(*t*), see Fig. 3). The ten deaths threshold was set to avoid incidental fluctuations that do not reflect the mortality trend of a certain country. Thresholds of five and 20 deaths led to similar results. We chose an absolute threshold rather than a relative threshold (i.e., number of death incidents normalized to population size) because the very initial dynamics of the disease is not expected to be strongly coupled with the population size. Furthermore, setting a relative threshold of one death per 1 million (or more) citizens is problematic for countries such as Iceland because its population size is smaller than 10^6^. However, setting a threshold of one death per 10^5^ citizens is problematic for countries such as Australia because such countries would never reach the starting threshold (at the date the data were collected, Australia had a mortality rate of 4 × 10^−6^ death cases per population size).

We fitted a linear regression model to these data (Fig. 4; excluding Japan), i.e., between *r* and the log *COVID-19 Mortality Probability* (denoted as *f*(*τ*)). This fitting resulted in the inferred model: log_10_ *f*(*τ*) = 0.04*τ* − 3.54 (slope 95% CI 0 05]). Let *τ*′ be an arbitrary response time point and let be the time with twice the number of deaths, i.e., *f*(*τ*″) = 2*f*(*τ*′). Therefore,

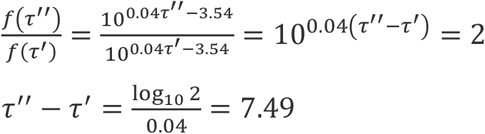

Resulting in a doubling time of 7.49 days with 95% *CI* [6.02,6 03].

### The offset between mortality growth rate and mobility data

The mortality growth rate measures the relative daily increase of fatalities and is defined by 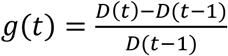. The optimal time offset, namely, the offset that yields the highest correlation between the declining trends of mobility *M*(*t*) and mortality growth rate *g*(*t*), is computed by *argmax*_Δt_{*corr*(*g*(*t* − Δ), *M*(*t*))} over all possible offsets, Δ (different offset examples are shown in Supplementary Fig. S3).

## Data Availability

All URLs for the data used in this study are available in the references section of this manuscript.

## Acknowledgements

G.L., S.A., O.A., K.H., and N.N. were supported in part by a fellowship from the Edmond J. Safra Center for Bioinformatics at Tel Aviv University. S.A. was partly supported by the Rothchild Caesarea Foundation. O.A. was partly supported by the Dalia and Eli Hurvits foundation. The authors wish to thank Itsik Pe’er for comments.

